# Impact of early postoperative ambulation on gait recovery after hip fracture surgery: A multicenter cohort study

**DOI:** 10.1101/2024.09.12.24313534

**Authors:** Keisuke Nakamura, Yasushi Kurobe, Keita Sue, Shinichi Sakurai, Tomohiro Sasaki, Shuhei Yamamoto, Naoko Ushiyama, Masahito Taga, Kimito Momose

## Abstract

**Objective:** This study aimed to investigate the effect of early postoperative ambulation on gait recovery at the initial postoperative week and at discharge after hip fracture surgery in older patients.

**Design:** Multicenter prospective cohort study.

**Setting and Participants:** The study included 882 patients aged ≥65 years from 10 acute hospitals in Japan.

**Methods:** Patients were divided into two groups according to the interval between surgery and first ambulation: early-ambulation (EA) group (initiation of ambulation on postoperative day 1 or 2) and late-ambulation (LA) group (initiation of ambulation on postoperative day 3 or later). The Functional Independence Measure (FIM) was assessed 1 day postoperatively, 1 week postoperatively, and at discharge. Independent walking regardless of use of walking aids was defined as walking FIM ≥5. Confounding variables were age, mobility and cognitive function before injury, medical history, fracture type, and waiting days for surgery.

Multivariate logistic regression analysis was conducted to determine whether EA affected independent walking at 1 week postoperatively and at discharge.

**Results:** The number of patients in the EA and LA groups was 292 (33.1%) and 590 (66.9%), respectively. The number of patients walking independently 1 week postoperatively and at discharge was 156 (17.7%) and 292 (33.1%), respectively. Multivariate logistic regression analysis revealed that EA was associated with independent walking at 1 week postoperatively (odds ratio [OR], 3.27; 95% confidence interval [CI], 2.17-4.94; *P* < .0001) and at discharge after adjusting for confounders (OR, 3.33; 95% CI, 2.38-4.69; *P* < .0001). EA was associated with the recovery to pre-injury walking status at discharge after adjusting for confounders (OR, 3.05; 95% CI, 1.59–5.93; *P* =.0009).

**Conclusion and Implications:** Early ambulation after hip fracture surgery has an impact on independent walking and recovery of pre-injury walking status at 1 week postoperatively and at discharge from acute hospitals in older patients.

## Introduction

The global incidence of hip fracture is estimated at 16.75 million annually, and it is expected to increase further [1, 2]. Hip fractures significantly impact mobility, leading to increased mortality, refracture rates, and the need for care [3, 4]. Previous studies have shown that approximately 20%-30% of patients who can walk pre-fracture fail to recover their pre-fracture walking status at 2 weeks or 6 months [5, 6]. The inability to walk at hospital discharge is an independent predictor of 1-year mortality [4]. Therefore, enhancing postoperative mobility is crucial for reducing mortality and mitigating long-term care and health-care expenses.

The guidelines for hip fracture management has recommended that patients undergo physiotherapy assessment and, unless medically or surgically contraindicated, mobilization on the day after surgery because early mobilization is associated with survival and recovery for patients after hip fracture [7–9]. The National Hip Fracture Database defines mobilization as the ability to sit or stand out of bed with or without help [7, 10]. Previous studies have shown that early mobilization leads to reduced mortality, reduced hospital stay, and increased ambulatory recovery [7, 11–13]. However, research on the impact of the early initiation of gait practice on gait recovery after surgery remains limited. In a previous randomized controlled trial study, those who started ambulation practice within 48 h postoperatively had higher gait reacquisition at 1 week postoperatively and were more likely to be discharged home than those who started ambulation practice later [14]. The impact of initiating early postoperative ambulation practice on gait recovery after the initial postoperative week remains uncertain.

This multicenter study aimed to investigate the effect of early ambulation on gait recovery after hip fracture surgery in the initial postoperative week and at discharge. We hypothesized that early postoperative ambulation (initiation of ambulation on postoperative day 1 or 2), would be associated with better gait recovery than late postoperative ambulation (initiation of ambulation on postoperative day 3 or later).

## Methods

### Study Design and Participants

This multicenter cohort study utilized the Nagano hip fracture database in Japan. This database contains information on the characteristics and rehabilitation outcomes of patients admitted with hip fracture, and between December 1, 2019, and July 31, 2023, data of 1613 surgically treated consecutive patients from 17 hospitals were submitted. These data were collected through a cloud electronic data capture system from each hospital. Patients who were ambulatory, aged 65 years or older before the fracture, had a femoral neck or trochanteric fracture, and admitted to an acute care hospital were included in the study.

Patients with postoperative weight-bearing restrictions were excluded from the study. A flowchart of the patient selection process is presented in Figure 1.

**Figure 1.**
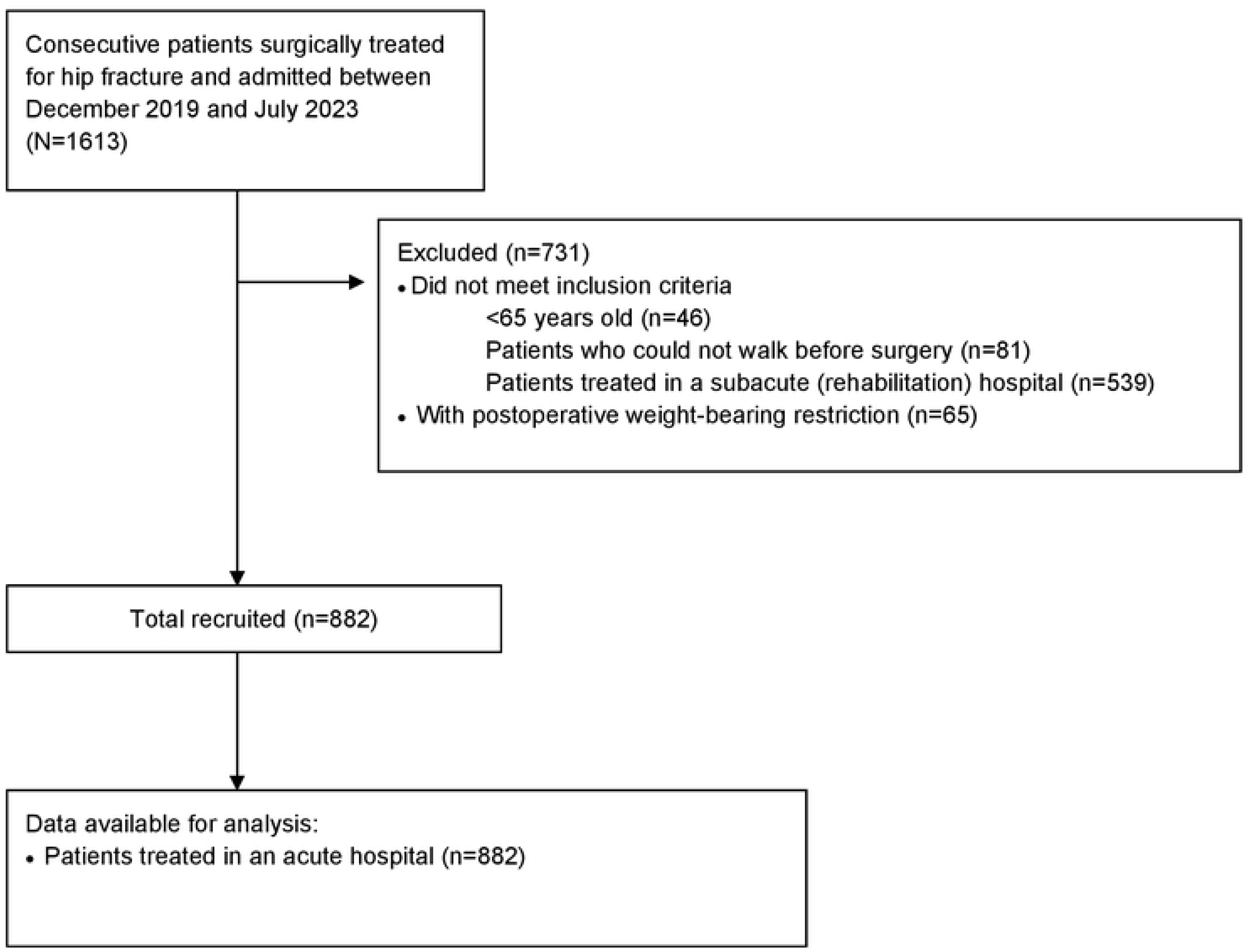
Patients’ recruitment and flow diagram.

All procedures were approved by the ethics committee of the School of Medicine, Shinshu University, on November 12, 2019 (protocol number 4541), and this study was performed according to the guidelines in the Declaration of Helsinki. Although this study did not require individual consent from participants, as it was an observational study and used anonymized data from normal medical practice, information on the conduct of the study was made public at each site. Participants were guaranteed the opportunity to opt out of the study at any time. Study details, including the objectives, inclusion and exclusion criteria, and primary outcome, were published in the publicly available University Hospital Information Network (UMIN-CTR, unique identifier: UMIN000054114.

### Measurement

The database contains the following information: patient background factors such as age, sex, body mass index, medical history (respiratory, cardiovascular, and neurological diseases), cognitive function before injury, mobility before injury, place of residence before admission, and residence after discharge, and medical factors such as fracture type, surgical procedure, complications (deep vein thrombosis, peroneal nerve palsy, infection, and falls), waiting days for surgery, waiting days for rehabilitation from admission, and rehabilitation factors (postoperative days to the start the ambulation and rehabilitation intervention time). Infections included pneumonia and urinary tract infections, and those diagnosed by a doctor were defined as having an infection. Cognitive function was assessed using the Degree of Daily Life Independence Score for People with Dementia (DDLIS-PD) [15, 16]. This assessment has seven scales and is widely used to evaluate dementia in Japan. In this study, cognitive impairment was defined as DDLIS-PD grade II, with independence by support with some hindrances, or higher grades. Functional Independence Measure (FIM) [17] at 1 day after surgery, 1 week postoperatively, and at discharge and walking status at discharge (e.g., walking without aids, with one-point cane, with walker and other aids) were assessed as treatment outcomes.

### Dependent Variables

The main outcome was independent walking regardless of the use of aids, defined as mobility (walking) with FIM grade ≥5, following the approach outlined by Fu et al [17]. A FIM score of ≥5 for walking indicates that the patient is able to walk with supervision or independently [18]. Walking ability was assessed 1 week postoperatively and at discharge. The secondary treatment outcome was walking recovery, which was defined as restoration of pre-fracture walking status and FIM grade ≥5. The other treatment outcomes were FIM score at discharge, walking status at discharge (ambulating without or with one-point cane, walker, or wheelchair), and days from operation to discharge.

### Independent Variables

The independent variable was early postoperative ambulation following hip fracture surgery. Patients were divided into two groups according to the interval between surgery and their first postoperative walk: the early-ambulation (EA) group (initiation of ambulation on postoperative day 1 or 2) and the late-ambulation (LA) group (initiation of ambulation on postoperative day 3 or later) [8, 14]. The date of ambulation initiation was defined as the date when a patient started ambulation regardless of the amount of the assistance or the use of walking aids.

We adjusted for confounding variables identified in previous studies, including age [19–23], walking status before injury [17, 21–25], cognitive impairment [12, 20–22, 24–26], preoperative medical history [18, 19, 21, 23] of respiratory, circulatory, and neurological diseases, fracture type [19, 21, 23, 24], and days from admission to surgery [6, 24], in the analysis of factors influencing postoperative walking independence.

### Statistical Analyses

Continuous variables are presented as median (25th and 75th percentile), whereas categorical variables are expressed as numbers and percentages. Pearson’s chi-squared test, Fisher’s exact test for count data, and Mann–Whitney U test were used to compare the rehabilitation factors and treatment outcomes between the EA and LA groups.

Multivariate logistic regression analysis was conducted to ascertain whether early postoperative ambulation initiation affected independent walking at 1 week postoperatively and at discharge. In this analysis, the dependent variable was whether the patient walked independently at 1 week postoperatively and at discharge, whereas the independent variable was early postoperative ambulation. All confounding factors were reduced to one composite characteristic by applying a propensity score. The propensity score, which reflected the likelihood of study participants being assigned to the EA or LA group, was used in logistic regression analysis as an independent variable [27]. Missing values were excluded because they represented <5% of the total [28].

As a sensitivity analysis to assess the influence of missing data on postoperative walking timing and recovery, we performed logistic regression analysis with multiple imputations of missing sites. To address missing data in the dataset, we performed multiple imputation using the “mice” package in R, version 4.3.2 (https://www.r-project.org), creating 20 complete datasets using methods such as predictive mean matching for continuous variables and logistic regression for categorical variables. Subsequently, logistic regression models were applied to each imputed dataset, and the results across imputed datasets were integrated using Rubin’s rules [29]. We further examined the potential impact of unknown confounders on the relationship between postoperative early first ambulation and independent walking by calculating the E-values (https://www.evalue-calculator.com/) [30, 31]. Statistical analyses were performed with R, version 4.3.2 (https://www.r-project.org). Statistical significance was set at *P* < .05.

## Results

A total of 882 patients with a median age (25^th^, 75^th^ percentiles) of 87 (81, 91) years from 10 acute hospitals were included, with 292 (33.1%) and 590 (66.9%) patients in the EA and LA groups, respectively. The patients’ demographic data are presented in Table 1. The median ages (25^th^, 75^th^ percentiles) of the EA and LA groups were 84 (79, 89) and 88 (83, 92) years, respectively; 78.8% and 46.6% walked without aids before the injury, and the percentages of cognitive impairment were 30.5% and 53.6%, respectively (Table 1).

**Table 1.**
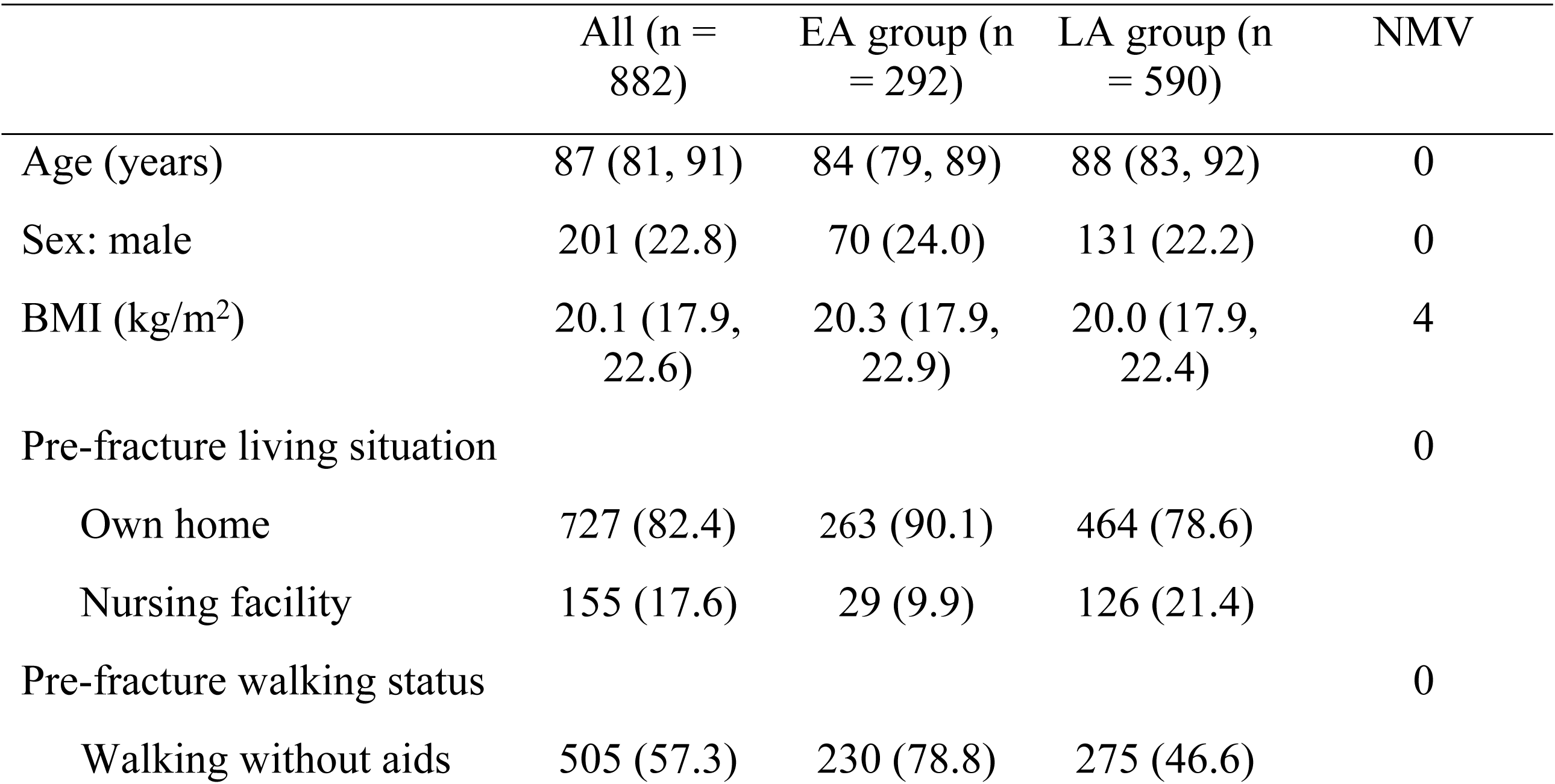
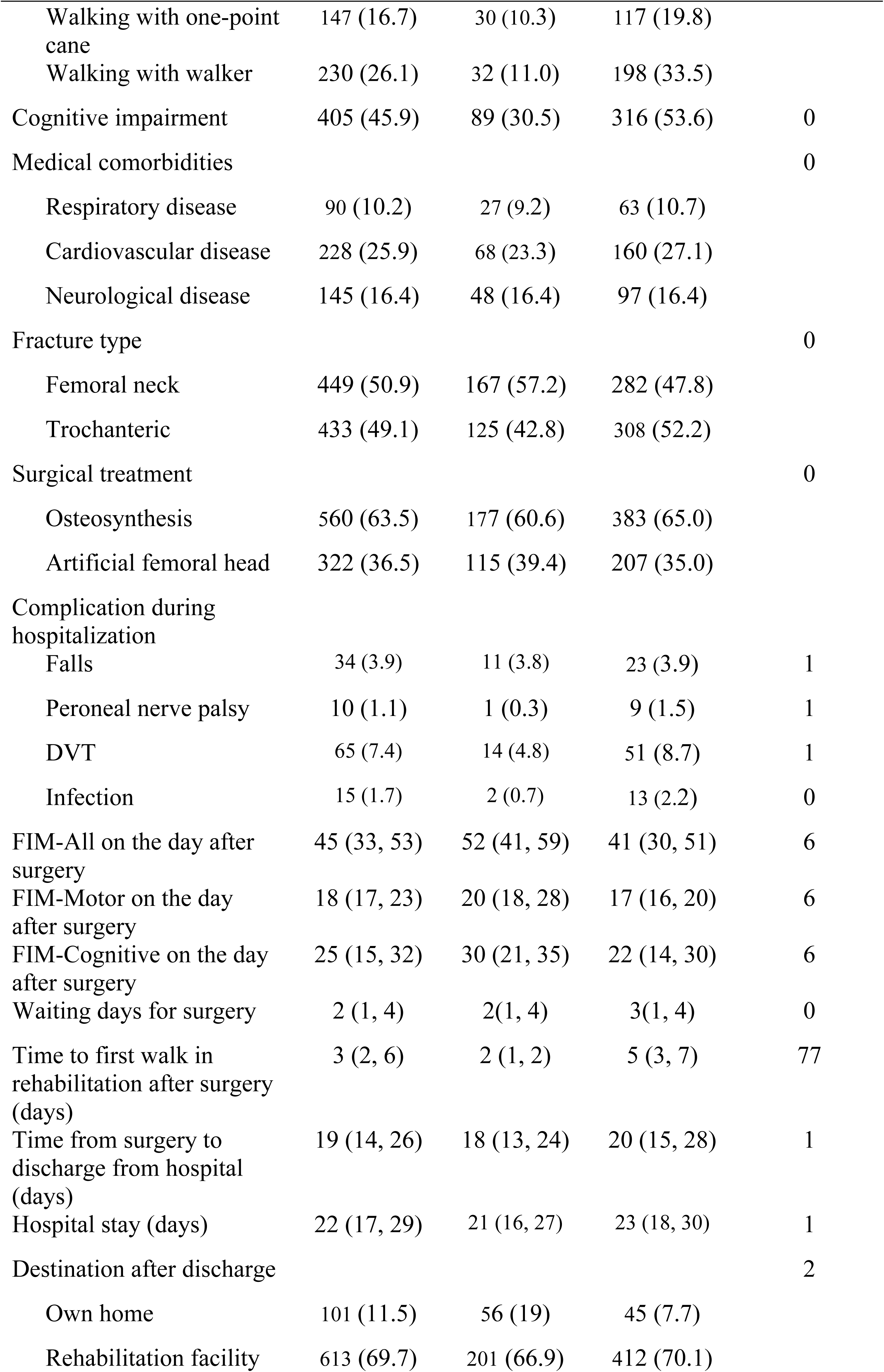
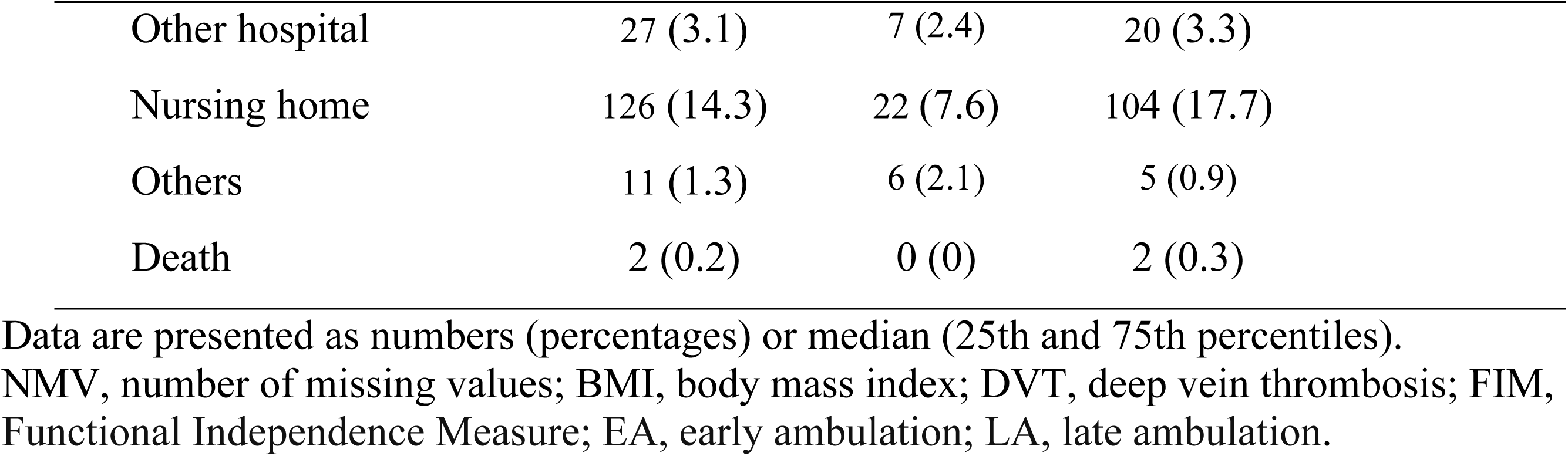
Patients’ demographic data.

|  | All (n = 882) | EA group (n = 292) | LA group (n = 590) | NMV |
| --- | --- | --- | --- | --- |
| Age (years) | 87 (81, 91) | 84 (79, 89) | 88 (83, 92) | 0 |
| Sex: male | 201 (22.8) | 70 (24.0) | 131 (22.2) | 0 |
| BMI (kg/m<sup>2</sup>) | 20.1 (17.9, 22.6) | 20.3 (17.9, 22.9) | 20.0 (17.9, 22.4) | 4 |
| Pre-fracture living situation |  |  |  | 0 |
| Own home | 727 (82.4) | 263 (90.1) | 464 (78.6) |  |
| Nursing facility | 155 (17.6) | 29 (9.9) | 126 (21.4) |  |
| Pre-fracture walking status |  |  |  | 0 |
| Walking without aids | 505 (57.3) | 230 (78.8) | 275 (46.6) |  |
| Walking with one-point cane | 147 (16.7) | 30 (10.3) | 117 (19.8) |  |
| Walking with walker | 230 (26.1) | 32 (11.0) | 198 (33.5) |  |
| Cognitive impairment | 405 (45.9) | 89 (30.5) | 316 (53.6) | 0 |
| Medical comorbidities |  |  |  | 0 |
| Respiratory disease | 90 (10.2) | 27 (9.2) | 63 (10.7) |  |
| Cardiovascular disease | 228 (25.9) | 68 (23.3) | 160 (27.1) |  |
| Neurological disease | 145 (16.4) | 48 (16.4) | 97 (16.4) |  |
| Fracture type |  |  |  | 0 |
| Femoral neck | 449 (50.9) | 167 (57.2) | 282 (47.8) |  |
| Trochanteric | 433 (49.1) | 125 (42.8) | 308 (52.2) |  |
| Surgical treatment |  |  |  | 0 |
| Osteosynthesis | 560 (63.5) | 177 (60.6) | 383 (65.0) |  |
| Artificial femoral head | 322 (36.5) | 115 (39.4) | 207 (35.0) |  |
| Complication during hospitalization |  |  |  |  |
| Falls | 34 (3.9) | 11 (3.8) | 23 (3.9) | 1 |
| Peroneal nerve palsy | 10 (1.1) | 1 (0.3) | 9 (1.5) | 1 |
| DVT | 65 (7.4) | 14 (4.8) | 51 (8.7) | 1 |
| Infection | 15 (1.7) | 2 (0.7) | 13 (2.2) | 0 |
| FIM-All on the day after surgery | 45 (33, 53) | 52 (41, 59) | 41 (30, 51) | 6 |
| FIM-Motor on the day after surgery | 18 (17, 23) | 20 (18, 28) | 17 (16, 20) | 6 |
| FIM-Cognitive on the day after surgery | 25 (15, 32) | 30 (21, 35) | 22 (14, 30) | 6 |
| Waiting days for surgery | 2 (1, 4) | 2(1, 4) | 3(1, 4) | 0 |
| Time to first walk in rehabilitation after surgery (days) | 3 (2, 6) | 2 (1, 2) | 5 (3, 7) | 77 |
| Time from surgery to discharge from hospital (days) | 19 (14, 26) | 18 (13, 24) | 20 (15, 28) | 1 |
| Hospital stay (days) | 22 (17, 29) | 21 (16, 27) | 23 (18, 30) | 1 |
| Destination after discharge |  |  |  | 2 |
| Own home | 101 (11.5) | 56 (19) | 45 (7.7) |  |
| Rehabilitation facility | 613 (69.7) | 201 (66.9) | 412 (70.1) |  |

|  |  |  |  |
| --- | --- | --- | --- |
| Other hospital | 27 (3.1) | 7 (2.4) | 20 (3.3) |
| Nursing home | 126 (14.3) | 22 (7.6) | 104 (17.7) |
| Others | 11 (1.3) | 6 (2.1) | 5 (0.9) |
| Death | 2 (0.2) | 0 (0) | 2 (0.3) |

Data are presented as numbers (percentages) or median (25th and 75th percentiles). NMV, number of missing values; BMI, body mass index; DVT, deep vein thrombosis; FIM, Functional Independence Measure; EA, early ambulation; LA, late ambulation.

The number of patients walking independently 1 week postoperatively and at discharge was 156 (17.7%) and 292 (33.1%), respectively. The proportion of patients walking independently at 1 week postoperatively and at discharge decreased with increasing duration to postoperative first ambulation (Supplementary Figure 1). The EA group had a significantly higher percentage of independent walking patients, defined as walking FIM grade ≥5 at 1 week postoperatively and at discharge, than the LA group (*P* < .0001) (Table 2). The EA group had significantly better treatment outcomes than the LA group, including walking status at discharge (*P* < .0001) and FIM at discharge (*P* < .0001) (Table 2).

**Table 2.** Treatment outcomes.

|  | EA group (n<br>= 292) | LA group (n<br>= 590) | P-value | NMV |
| --- | --- | --- | --- | --- |
| Walking status at discharge |  |  | <.0001 | 13 |
| Walking without aids | 13 (4.5) | 6 (1.0) |  |  |
| Walking with one-point cane | 48 (16.5) | 25 (4.3) |  |  |
| Walking with walker | 110 (37.8) | 90 (15.6) |  |  |
| Wheelchair | 120 (41.2) | 455 (78.7) |  |  |
| Death | 0 (0) | 2 (0.3) |  |  |
| FIM-All at discharge | 84 (67, 100) | 61 (43, 82) | <.0001 | 18 |
| FIM-Motor at discharge | 55 (45, 68) | 38 (24, 52) | <.0001 | 18 |
| FIM-Cognitive at discharge | 31 (23, 35) | 23 (15, 31) | <.0001 | 18 |
| Walking FIM at 1 week postoperatively $\geq 5$ | 103 (36.7) | 53 (9.2) | <.0001 | 28 |
| Walking FIM at discharge $\geq 5$ | 171 (58.8) | 121 (21.0) | <.0001 | 15 |
| Recovery to pre-injury walking status | 25 (8.6) | 21 (3.6) | .002 | 15 |

Data are presented as numbers (percentages) or median (25th and 75th percentiles). NMV, number of missing values; FIM, Functional Independence Measure; EA, early ambulation; LA, late ambulation.

Based on multivariate logistic regression analysis, EA (≤2 days from surgery to first ambulation) was associated with independent walking at 1 week postoperatively and at discharge after adjusting for confounders, with odds ratios (ORs) of 3.27 (95% confidence interval [CI], 2.17-4.94) and 3.33 (95% CI, 2.38-4.69), respectively (Table 3). Furthermore, EA was associated with the recovery to pre-injury walking status at discharge after adjusting for confounders, with OR of 3.05 (95% CI, 1.59-5.93) (Table 4).

**Table 3.** Odds ratios for walking independence at 1 week postoperatively and at discharge from acute hospitals following early postoperative initiation of ambulation in older patients who underwent hip fracture surgery.

| Variables | Patients | Walking<br>independently<br>rate | Adjusted<br>odds ratio* | 95% CI |  | P-value | Adjusted<br>odds ratio* | 95% CI |  | P-value |
| --- | --- | --- | --- | --- | --- | --- | --- | --- | --- | --- |
|  |  |  |  | Lower | Upper |  |  | Lower | Upper |  |
| At 1 week postoperatively |  |  | Completed data (n = 858) |  |  |  | Multiple imputation data |  |  |  |
| LA (>2 days from surgery to first walk) | 590 (66.9) | 53 (9.2) | 1.00 [ref] |  |  |  | 1.00 [ref] |  |  |  |
| EA (≤2 days from surgery to first walk) | 292 (33.1) | 103 (36.7) | 3.27 | 2.17 | 4.94 | <.0001 | 3.21 | 2.14 | 4.83 | <.0001 |
| At discharge from acute hospital |  |  | Completed data (n = 867) |  |  |  | Multiple imputation data |  |  |  |
| LA (>2 days from surgery to first walk) | 590 (66.9) | 121 (21.0) | 1.00 [ref] |  |  |  | 1.00 [ref] |  |  |  |
| EA (≤2 days from surgery to first walk) | 292 (33.1) | 171 (58.8) | 3.33 | 2.38 | 4.69 | <.0001 | 3.28 | 2.34 | 4.59 | <.0001 |

CI, confidence interval; ref, reference; EA, early ambulation; LA, late ambulation.
\*Applying the propensity score calculated using a model for predicting the postoperative ambulation timing, whether early or late ambulation, considering the confounding variables: age, preoperative walking ability, cognitive impairment, preoperative medical history of respiratory, circulatory, and neurological diseases, fracture type, and preoperative waiting period.

**Table 4.** Odds ratios for the recovery to pre-injury walking status following early postoperative initiation of ambulation in older patients who underwent hip fracture surgery.

| Variables | Patients,<br>n (%) | Walking<br>recovery**<br>rate, n (%) | Adjusted<br>Odds Ratio* | 95%CI |  | *p*-value | Adjusted<br>Odds Ratio* | 95%CI |  | *p*-value |
| --- | --- | --- | --- | --- | --- | --- | --- | --- | --- | --- |
|  |  |  |  | Lower | Upper |  |  | Lower | Upper |  |
| Recovery to pre-injury walking status<br>at discharge | Completed data (N=867) |  |  |  |  |  | Multiple imputation data |  |  |  |
| LA (>2 days from surgery to first<br>walk) | 590 (66.9%) | 21 (3.6) | 1.00 [ref] |  |  |  | 1.00 [ref] |  |  |  |
| EA (≤ 2 days from surgery to first<br>walk) | 292 (33.1%) | 25 (8.6) | 3.05 | 1.59 | 5.93 | 0.0009 | 2.83 | 1.47 | 5.44 | 0.001 |

CI, confidence interval; ref, reference; EA, early ambulation; LA, late ambulation
\* Applying propensity score calculated using a model to predict the postoperative ambulation timing, whether early or late ambulation, considering the confounding variables: age, preoperative walking ability, cognitive impairment, preoperative medical history of respiratory, circulatory, and neurological diseases, fracture type, and preoperative waiting period.
\*\* Walking recovery was defined as restoration of pre-fracture walking status and FIM grade ≥5.

Sensitivity analysis showed similar results for models with multiple assignments of missing values for the completed data models. Subsequently, to evaluate the influence of potential unmeasured confounders on the relationship between postoperative early first walk and independent walking at 1 week postoperatively and at discharge, we calculated the E-values, which were 3.02 and 3.05, respectively.

## Discussion

This multicenter cohort study investigated the effect of early postoperative ambulation on gait recovery at the initial postoperative week and at discharge after hip fracture surgery in older patients, and the results showed that the initiation of ambulation within 2 postoperative days after hip fracture surgery was associated with independent walking at 1 week postoperatively and at discharge. Furthermore, early postoperative ambulation may influence the recovery of walking to pre-injury status. Other effects of early initiation of ambulation were reduced length of hospital stay, discharge home, and improved ability to perform activities of daily living.

The effect of early ambulation initiation in the present study supports the results of previous research [14, 32]. Oldmeadow et al. conducted a study to evaluate the effects of early assisted ambulation, defined as the first walk on postoperative day 1 or 2, compared with delayed assisted ambulation, defined as the first walk on postoperative day 3 or 4, in an acute hospital setting. The results showed that at 1 week after surgery, the early ambulation group required less assistance for transfers and ambulation and walked farther [14]. In the present study, the results showed a longer-term effect on gait recovery at discharge (approximately 3 weeks postoperatively) by early ambulation initiation, similar to the results of previous studies [14]. The present study showed that early postoperative ambulation initiation was effective in independent walking and recovery of walking status before injury, even after adjusting for the confounding factors mentioned in previous studies, such as age [19–23], pre-fracture mobility [17, 21–25], cognitive function [12, 20–22, 24–26], comorbidity disease [17, 20, 22], fracture type [19, 21, 23, 24], and days from admission to surgery [6, 24]. Oldmeadow et al. stated that cardiovascular stability is a major determinant of successful early ambulation after hip fracture surgery, and thus, the present study used the presence of cardiovascular disease as a confounding factor [14]. Although the association between early postoperative ambulation initiation and gait reacquisition in this study is unclear, it is possible that early ambulation initiation prevented disuse and increased the amount of ambulation practice prior to discharge compared with late ambulation initiation, which may have resulted in the reacquisition of independent walking. Shimizu et al. and Marsault et al. reported that higher physical activity during hospitalization for hip fracture patients was associated with higher ability to perform activities of daily living at discharge [33, 34]. However, these studies assessed physical activity for only a short period of 3 days to approximately 1 week [33, 34]; therefore, future studies are needed to investigate the relationships between postoperative physical activity and independent walking.

There are several unmeasured confounding factors in this study [35], such as pain [36], delirium [37], nutritional status [6, 25, 38], and inflammation [6, 32], which affect early postoperative ambulation initiation and gait reacquisition according to previous studies.

Postoperative hip fracture-related pain was associated with a trochanteric fracture [36]; thus, we can potentially take into account the impact of pain on gait reacquisition by adjusting for the confounding factor of a trochanteric fracture. With regard to delirium, a previous study reported a strong association between delirium and cognitive decline [39]. The present study adjusted for cognitive decline as a confounding factor, which is believed to consider the influence of delirium. In the present study, to evaluate the influence of potential unmeasured confounders, we calculated the E-values, and the resulting E-values, 3.02 and 3.05, exceeded the odds ratio of unobserved confounders independently affecting the association between early ambulation and postoperative ambulation, suggesting a minimal effect of unknown or unmeasured confounders on this association.

This study has some limitations. First, the reasons for not being able to start walking early postoperatively were unclear. Oldmeadow et al. reported that early postoperative ambulation initiation failed when patients were medically unstable [14]. Other studies have found that reasons for delaying mobilization include pain, fatigue, and habitual cognitive status [36].

Second, the effect of early postoperative ambulation initiation on long-term gait reacquisition 6 months or 1 year later is unknown. Finally, in this study, walking independence was defined as walking FIM ≥5, which is the ability to walk independently with or without aids for at least 15 m, which indicates the least ability of independent indoor walking [18], similar to the findings of Fu et al. Some walking status assessments were used in the previous studies, such as FIM [17], the Iowa Level of Assistance scale [14, 32], the Cumulated Ambulation Score [12], and walking status according to use of walking aids [6, 21]. In the future, more applied assessments of walking ability will need to use outcomes other than the FIM.

The strengths of this study include the large population with a multicenter cohort, the prospective design, and the systematic data collection, which eliminate single-center bias and may increase the generalizability of the results. Furthermore, the selected outcomes are of great importance in clinical practice. In addition, the independent variables used in this study included the most important factors known to be associated with the outcomes. This study reflects daily clinical practice in the acute hospital.

## Conclusions and Implications

The study results indicate that initiating ambulation within 2 postoperative days after hip fracture surgery affects independent walking at postoperative week 1 and at discharge.

Furthermore, early postoperative ambulation may influence the recovery of walking to pre-injury status. However, the reasons for delayed ambulation postoperatively were unclear.

Future investigation into the reasons for delayed ambulation could lead to a multidisciplinary team approach to encourage early ambulation within 2 days of surgery.

## Data Availability

The data that support the findings of this study are available from the corresponding author (Keisuke Nakamura) upon reasonable request.

## Acknowledgements

We thank all the participants who cooperated in our study as well as the rehabilitation staff in the cooperating hospitals for their assistance with data collection for the study.

